# Transmission blocking activity of low dose tafenoquine in healthy volunteers experimentally infected with *Plasmodium falciparum*

**DOI:** 10.1101/2022.03.15.22272229

**Authors:** Rebecca Webster, Hayley Mitchell, Jenny M. Peters, Juanita Heunis, Brighid O’Neill, Jeremy Gower, Sean Lynch, Helen Jennings, Fiona H. Amante, Stacey Llewellyn, Louise Marquart, Adam J. Potter, Geoffrey W. Birrell, Michael D. Edstein, G. Dennis Shanks, James S. McCarthy, Bridget E. Barber

**Author notes:** **Corresponding author:** Dr. Bridget E. Barber. QIMR Berghofer Medical Research Institute, 300 Herston Road, Herston 4006, Australia.

## Abstract

**Background:** Blocking the transmission of parasites from humans to mosquitoes is a key component of malaria control. Tafenoquine exhibits activity against all stages of the malaria parasite and may have utility as a transmission blocking agent. We aimed to characterize the transmission blocking activity of low dose tafenoquine.

**Methods:** Healthy adults were inoculated with *P. falciparum* 3D7-infected erythrocytes on day 0. Piperaquine was administered on days 9 and 11 to clear asexual parasitemia while allowing gametocyte development. A single 50 mg oral dose of tafenoquine was administered on day 25. Transmission was determined by enriched membrane feeding assays pre-dose and at 1, 4 and 7 days post-dose. Artemether-lumefantrine was administered following the final assay. Outcomes were the reduction in mosquito infection and gametocytemia post-tafenoquine, and safety parameters.

**Results:** Six participants were enrolled, and all were infective to mosquitoes pre-tafenoquine, with a median 86% (range: 22–98) of mosquitoes positive for oocysts and 57% (range: 4–92) positive for sporozoites. By day 4 post-tafenoquine, the oocyst and sporozoite positivity rate had reduced by a median 35% (IQR: 16–46) and 52% (IQR: 40–62), respectively, and by day 7, 81% (IQR 36–92) and 77% (IQR 52–98), respectively. The decline in gametocyte density post-tafenoquine was not significant. No significant participant safety concerns were identified.

**Conclusion:** Low dose tafenoquine reduces *P. falciparum* transmission to mosquitoes, with a delay in effect.

**Trial registration:** Australian New Zealand Clinical Trials Registry (ACTRN12620000995976).

**Funding:** QIMR Berghofer Medical Research Institute.

## INTRODUCTION

Despite advances in the development and implementation of tools to prevent and treat malaria since the turn of the century, worldwide malaria morbidity and mortality remains unacceptably high (1). The emergence of partial resistance to the first line artemisinin antimalarial compounds in Southeast Asia, and more recently in East Africa, is of particular concern (2). It is likely that further progress in reducing the burden of malaria and achieving the ambitious goal of malaria eradication in the 21^st^ century will require a multifaceted approach involving antimalarial chemotherapy, vaccination, and vector control.

The importance of targeting malaria transmission as an intervention strategy is highlighted by the fact that the distribution of insecticide-treated bed nets was the largest contributor to the declining prevalence of *P. falciparum* infections in Africa between 2000 and 2015 (3). Blocking the transmission of parasites from humans to mosquitoes using antimalarial chemotherapy represents another strategy for reducing the burden of malaria and slowing the spread of resistance to antimalarials. Transmission is mediated by circulating, sexually committed, mature male and female gametocytes. These gametocytes rapidly differentiate into gametes upon uptake in a blood meal by the female *Anopheles* mosquito. Following fertilization, the motile ookinete penetrates the midgut epithelium followed by oocyst development, sporogony and migration of sporozoites to the salivary glands (4). Antimalarial drugs used to treat clinical malaria by clearing asexual blood stage parasitemia typically demonstrate lower activity towards mature transmissible gametocytes due to their arrested state of cellular development (5).

The 8-aminoquinolone primaquine is currently the only drug recommended for blocking transmission of malaria (6); it is highly effective in clearing mature gametocytes and impairing the development of mosquito stage parasites after the ingestion of the blood meal (7, 8). However, the use of primaquine is limited by the hemolysis it causes in individuals with deficient activity of the glycolytic enzyme glucose-6-phosphate dehydrogenase (G6PD), a common genetic disorder in malaria-endemic populations (9). The World Health Organization (WHO) thus recommends that primaquine only be used at a low dose (0.25 mg/kg single dose) when treating individuals of unknown G6PD status (6). Another limitation of primaquine is its short elimination half-life (plasma half-life approximately 7 hours) (10), which limits the duration of its pharmacodynamic activity both for transmission blocking and as a drug with pre-erythrocytic activity against incubating liver stage parasites. This particularly limits its utility for mass drug administration or for seasonal malaria chemoprophylaxis.

Tafenoquine is a long acting analogue of primaquine that has been recently approved for malaria prophylaxis and for radical cure of *P. vivax* malaria (11). The major advantage of tafenoquine over primaquine is its considerably longer plasma elimination half-life (approximately 15 days) (12, 13) which allows for single dose (radical cure) or weekly administration for prophylaxis, thus reducing adherence issues commonly associated with primaquine dosing regimens (14). Although not currently approved for transmission blocking, the potential utility of tafenoquine for such is evident given its gametocytocidal activity *in vitro* (15) and transmission blocking activity in murine models (16). Like primaquine, tafenoquine can cause severe hemolytic anemia in G6PD-deficient individuals at currently recommended doses (17); however, as with primaquine, it is possible that a sufficient safety margin may be present for its therapeutic use in G6PD-deficient individuals (18). The ability of low dose tafenoquine to block malaria transmission is therefore worthy of investigation.

Volunteer infection studies (VIS) using the induced blood stage malaria (IBSM) model have been successfully utilized to characterize the antimalarial activity of several compounds in use or undergoing clinical development (19-23). These studies involve intravenous inoculation of healthy adult volunteers with blood stage parasites followed by dosing with the test antimalarial when a predefined parasitemia threshold is reached. IBSM studies have largely focused on charactering activity against blood stage asexual parasites. However, more recently we have demonstrated the potential utility of the model in assessing transmission blocking interventions (24). The IBSM-transmission model utilizes the antimalarial drug piperaquine to clear asexual parasites. As piperaquine lacks activity against gametocytes (25), the effect of transmission blocking interventions can be assessed subsequent to cure of asexual parasitemia with piperaquine when mature transmissible gametocytes re-enter circulation.

Here we report the results of a clinical trial to characterize the transmission blocking activity of a low single oral dose of tafenoquine (50 mg) in healthy volunteers using the IBSM-transmission model.

## RESULTS

### Participants

The study was conducted from 15 April 2021 to 23 June 2021. A total of 19 volunteers were assessed for eligibility, with 13 volunteers excluded for not meeting the inclusion/exclusion criteria (*n*=9) or due to the volunteers’ decision not to take part in the study (*n*=4). A total of 6 healthy, malaria naïve participants were enrolled and inoculated intravenously with *P. falciparum* 3D7 infected erythrocytes on day 0. Participants (3 male and 3 female) were aged between 21 and 55 years; 5 participants self-selected their race as White and one participant as Australian Aboriginal or Torres Strait Islander (Table 1). All participants were orally dosed with piperaquine on day 9 (480 mg) and day 11 (960 mg), and tafenoquine on day 25 (50 mg). All participants completed the study and were included in the analysis of study endpoints.

**Table 1.** Demographic profile of participants (*n*=6).

|  | Participant 1 | Participant 2 | Participant 3 | Participant 4 | Participant 5 | Participant 6 |
| --- | --- | --- | --- | --- | --- | --- |
| Age range (years) | 21-25 | 31-35 | 51-55 | 21-25 | 26-30 | 31-35 |
| Sex | Female | Male | Male | Female | Female | Male |
| Race | White | White | White | Australian Aboriginal or<br>Torres Strait Islander | White | White |
| Weight (kg) | 61 | 105 | 85 | 57 | 77 | 78 |
| Body mass index (kg/m<sup>2</sup>) | 24 | 32 | 30 | 21 | 28 | 27 |

### Parasitemia

The progression of parasitemia following intravenous inoculation with blood stage parasites (Figure 1A) was consistent between participants, with a median parasitemia of 12568 parasites/mL (range 4318–82140) recorded on day 9 prior to the commencement of piperaquine treatment. Piperaquine successfully cleared asexual parasitemia in all participants, while mature gametocytes were observed re-entering circulation from approximately day The absence of asexual ring stage parasites from day 16 was confirmed using qRT-PCR targeting the *SBP1* transcript which is specific for ring stage parasites, while the presence of both male and female gametocytes was confirmed by qRT-PCR targeting *pfMGET* and *pfs25* transcripts, respectively (Figure S1, supplementary file page 2). The median gametocyte density (as determined by 18S qPCR) at the time of dosing with 50 mg tafenoquine on day 25 was 234 gametocytes/mL (range 95–3234). The median 138 within-participant percent reduction in gametocyte density was not statistically significant at any time-point following tafenoquine dosing (Table 2). Similarly, the change in the ratio of *pfMGET* to *pfs25* transcripts from day 25 (pre-tafenoquine) to post-treatment time points (4 and 7 days post-dosing) was not significant (Table S1, supplementary file page 3).

**Figure 1.**
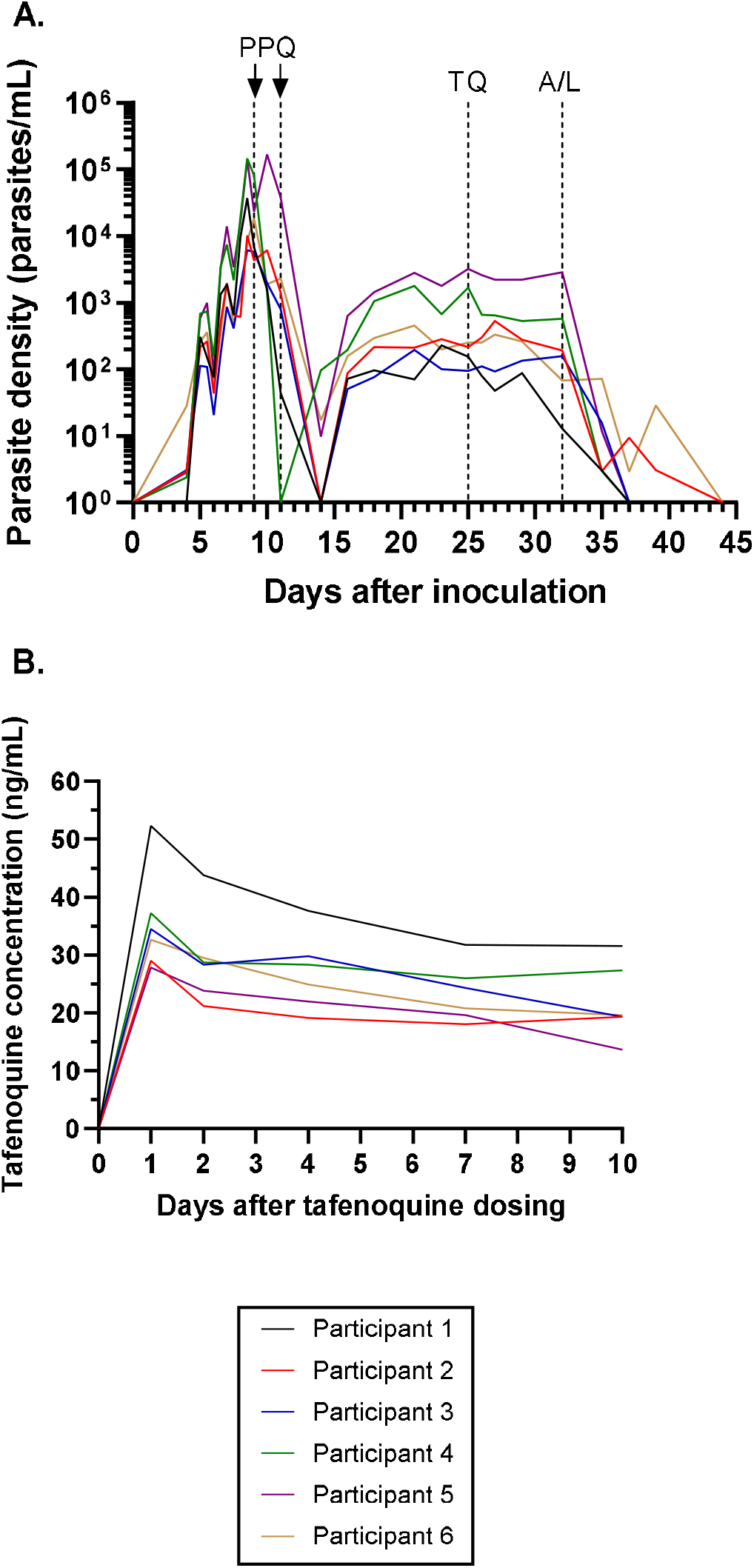
Individual participant parasitemia-time profiles (A) and tafenoquine plasma concentration-time profiles (B). Participants were inoculated intravenously with *P. falciparum*-infected erythrocytes on day 0 and were administered an oral dose of piperaquine (PPQ) on day 9 (480 mg) and day 11 (960 mg) to clear asexual parasitemia while allowing transmissible gametocytes to mature. A single oral dose of 50 mg tafenoquine (TQ) was administered on day 25. Definitive antimalarial treatment with a standard course of artemether-lumefantrine (A/L) was initiated on day 32. Parasitemia was measured using qPCR targeting the gene encoding *P. falciparum*18S rRNA. Tafenoquine plasma concentrations were measured using liquid chromatography tandem mass spectrometry.

**Table 2.** Change in gametocyte density from pre-tafenoquine dosing to post-treatment time points

| Time-point (days relative to tafenoquine dosing) | *N* | Median gametocyte density (range) | Median % reduction (IQR) | *P*-value* |
| --- | --- | --- | --- | --- |
| 0 (pre-tafenoquine) | 6 | 234 (95 – 3236) |  |  |
| 1 | 6 | 275 (79 – 2570) | 10 (-17 – 48) | 0.46 |
| 4 | 6 | 269 (89 – 2239) | 14 (-29 – 42) | 0.46 |
| 7 | 6 | 174 (13 – 2884) | 38 (10 – 73) | 0.17 |

Gametocyte density per mL was measured using 18S qPCR. Median % reduction is the median within participant percentage reduction in gametocyte density relative to the pre-tafenoquine time point (baseline). IQR: interquartile range. \*Reduction in gametocyte density was assessed using a Wilcoxon signed-rank test.

At day 26, the mean (range) plasma tafenoquine concentration was 35.6 ng/mL (27.8 to 52.3) at about 24 hours after the 50 mg dose of tafenoquine, and concentrations remained ≥18 ng/mL over the course of the 7 day period during which transmission was assessed (Figure 1B). Definitive antimalarial treatment with a standard 3-day course of artemether-lumefantrine was initiated on day 32 (7 days post tafenoquine dosing) for all participants. Two participants (participant 2 and participant 6) were administered a single dose of 45 mg primaquine after artemether-lumefrantrine treatment (day 44) to clear residual gametocytes. All participants were confirmed to be aparasitemic by qPCR prior to the conclusion of the study (Figure 1A).

### Transmission

Transmission was measured by detection of oocysts and sporozoites in mosquito midgut and head/thorax dissections, respectively, by 18S qPCR. Microscopy was also performed on dissected mosquito midguts stained with mercurochrome for visual confirmation of oocysts. The oocyst detection rate by 18S qPCR and microscopy was strongly correlated (r_rm_=0.95 [95% CI: 0.87 – 0.98], *P*<0.001), and the 18S qPCR results are reported henceforth.

All 6 participants were infective to mosquitoes (at both the oocyst and sporozoite level) prior to tafenoquine dosing (Figure 2A/B). The intensity of mosquito infection was high, with a median 86% (range 22 – 98) of mosquitoes positive for oocysts, and 57% (range 4 – 92) positive for sporozoites (Table 2). All 6 participants remained infective to mosquitoes (at both the oocyst and sporozoite levels) one day after tafenoquine dosing (Figure 2A/B), and no significant change in the intensity of mosquito infection was evident at this time point (Table 2). Most participants remained infective to mosquitoes at all time-points following tafenoquine dosing, with only one participant not infective to mosquitoes at the oocyst level on day 4 and day 7 post-dosing, and another not infective at the sporozoites level at day 7 (*P*=0.32 [McNemar’s test] for decline in proportion of participants infective to mosquitoes at either oocyst or sporozoite levels at day 7 post dosing). However, a significant decline in the intensity of mosquito infection was detected at days 4 and 7 post-dosing at both the oocyst and sporozoite levels (Table 2).

**Figure 2.**
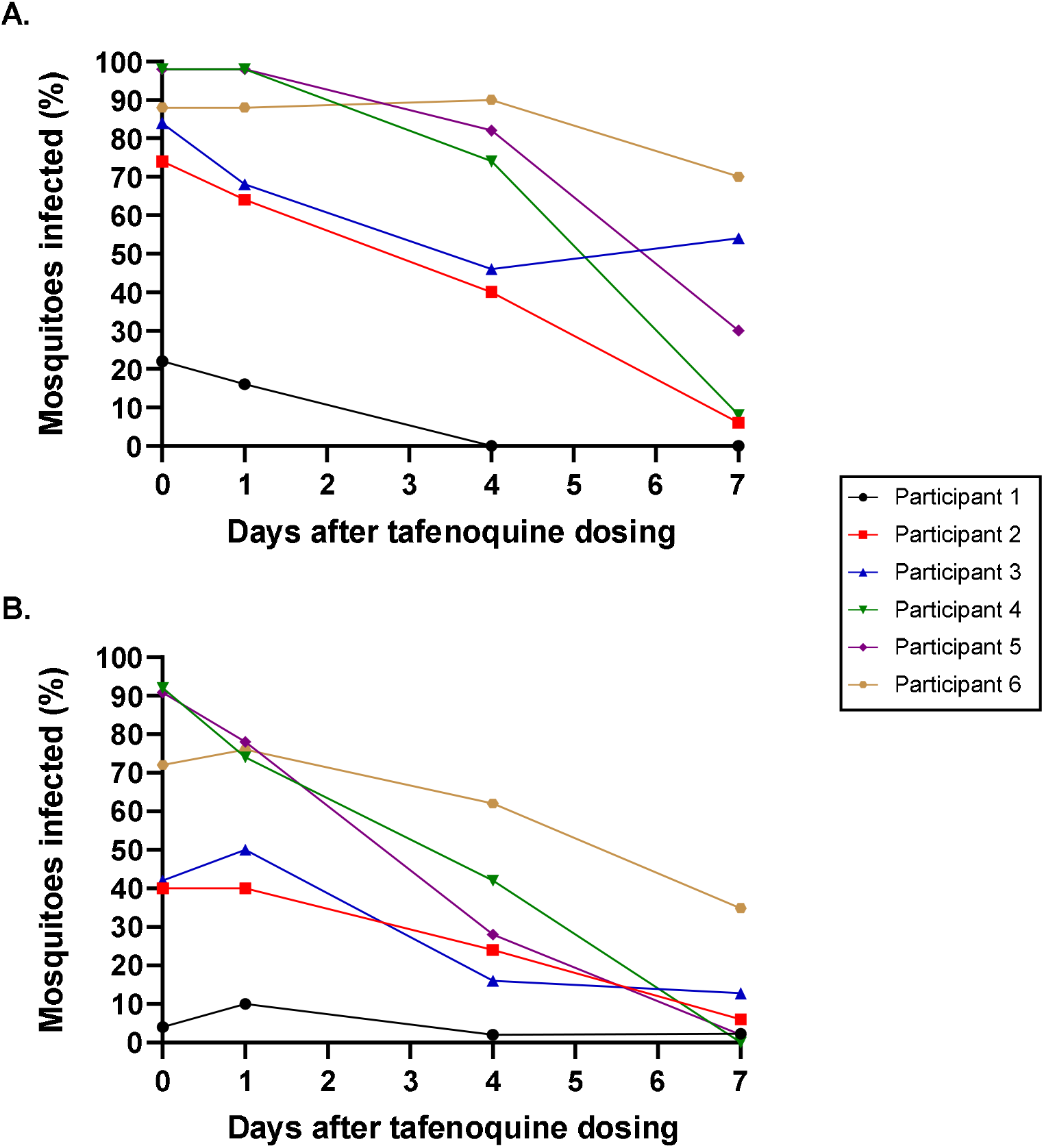
Individual participant mosquito infectivity rates at the oocyst (A) and sporozoite (B) levels. Blood samples were collected prior to and after administration of a single oral dose of 50 mg tafenoquine and transmission to mosquitoes was determined using an enriched membrane feeding assay. The presence of oocysts in mosquito midgut dissections (*n*=50 for each participant at each time point) and sporozoites in mosquito head/thorax dissections (*n*=42-50 for each participant at each time point) were detected using qPCR targeting the gene encoding *P. falciparum* 18S rRNA. The sporozoite and oocyst positivity rates were highly correlated (r_rm_=0.92 [95% CI: 0.79 – 0.97], p<0.001).

**Table 2.** Change in the intensity of mosquito oocyst and sporozoite infection from pre-tafenoquine dosing to post-treatment time points.

| Time-point<br>(days<br>relative to<br>tafenoquine<br>dosing) | *N* | Median %<br>infected<br>mosquitoes<br>(range) | Median %<br>reduction<br>(IQR) | *P*-<br>value* | Median %<br>infected<br>mosquitoes<br>(range) | Median %<br>reduction<br>(IQR) | *P*-value* |
| --- | --- | --- | --- | --- | --- | --- | --- |
|  |  | Oocysts |  |  | Sporozoites |  |  |
| 0 (pre-tafenoquine) | 6 | 86 (22 – 98) |  |  | 57 (4 – 92) |  |  |
| 1 | 6 | 78 (16 – 98) | 7 (0 – 19) | 0.087 | 62 (10 – 78) | -3 (-19 – 14) | 0.67 |
| 4 | 6 | 60 (0 – 90) | 35 (16 – 46) | 0.046 | 26 (2 – 62) | 52 (40 – 62) | 0.028 |
| 7 | 6 | 19 (0 – 70) | 81 (36 – 92) | 0.028 | 4 (0 – 35) | 77 (52 – 98) | 0.028 |

Median % reduction is the median within participant percentage reduction in the proportion of infected mosquitoes relative to the pre-tafenoquine dosing time point (baseline). \*Reduction in intensity of mosquito infection was assessed using a Wilcoxon signed-rank test.

### Predictors of infectivity to mosquitoes

At the pre-tafenoquine dosing time-point (day 25), infectivity to mosquitoes at the oocyst level was positively correlated with the level of gametocytemia at this time point, as measured by 18S qPCR (r=0.81, *P*=0.050), with a positive trend also noted at the sporozoite level (r=0.77, *P*=0.072). No correlation was observed between infectivity at day 25 and the ratio of *pfMGET* to *pfS25* transcripts at this time point (oocysts: r=0.20, p=0.70; sporozoites: r=0.14, p=0.79). Similarly, no correlation was observed between infectivity at day 25 and peak parasitemia at the time of piperaquine dosing (oocysts: r=0.58, p=0.23; sporozoites: r=0.54, p=0.27), or the area under the parasitemia-time curve prior to piperaquine dosing (oocysts: r=0.58, *P*=0.23; sporozoites: r=0.60, *P*=0.21).

Interestingly, in exploring possible explanations for why one participant may have had a lower intensity of mosquito infectivity at baseline compared to the other participants (Participant 1, Figure 2), we noted that this participant had a significantly elevated C reactive protein concentration (CRP, 46 mg/L) compared to the other participants (16 mg/L for Participant 2, 5 mg/L for Participants 3 and <5 mg/L for Participants 4 – 6). Replacing <5 mg/L with 2.5 mg/L, although numbers are small, a strong inverse correlation between CRP and infectivity was noted at both the sporozoite (r=-0.94, *P*=0.005) and the oocyst levels (r=-0.95, *P*=0.003).

### Safety and tolerability

A total of 63 adverse events (AEs) were reported in this study, the majority of which were mild signs and symptoms of malaria (Table 3 and Table S1, supplementary file, page 4). No AEs met the protocol-defined criteria of a serious adverse event (SAE), resulted in a participant discontinuing the study, or were considered related to dosing with tafenoquine. There were 14 AEs of moderate intensity and one AE of severe intensity. The severe AE was a low lymphocyte count (0.4×10^9^/L, normal range 1.1 - 4×10^9^/L) on day 9 (first piperaquine dose). The event spontaneously resolved without medical intervention over the course of one week (lymphocyte count 1.4×10^9^/L on day 16). A decreased white blood cell count (lymphocytes, neutrophils, and/or a composite decrease in leukocytes) was observed in four participants around the time of piperaquine dosing. Other than the severe (grade 3) decrease in lymphocyte count, the decrease in total white blood cell counts were moderate (grade 2) in intensity and spontaneously resolved over the course of a few days. All participants experienced a mild fall in hemoglobin, with a median fall of 21.5 g/L (range 15 – 36) from pre-inoculation to hemoglobin nadir (Figure S2, supplementary file page 3). Hemoglobin declines of >20 g/L occurred in 3 participants and were recorded as mild AEs; these events were considered unrelated to tafenoquine since the onset occurred prior to dosing. No clinically significant abnormalities in other laboratory parameters were recorded for any participant. Monitoring of vital signs revealed mild pyrexia in 2 participants and mild tachycardia in one participant. No clinically significant abnormalities were recorded for electrocardiogram (ECG) parameters.

**Table 3.** Summary of adverse events.

| Adverse event category<sup>a</sup> | Participant 1 | Participant 2 | Participant 3 | Participant 4 | Participant 5 | Participant 6 | Total<br>N=6 |
| --- | --- | --- | --- | --- | --- | --- | --- |
| Any adverse event | 9 | 6 | 7 | 17 | 21 | 3 | 63 |
| Adverse event of moderate intensity<br>(grade 2)<sup>b</sup> | 2 | 1 | 1 | 2 | 8 | 0 | 14 |
| Adverse event of severe intensity<br>(grade 3)<sup>b</sup> | 0 | 0 | 0 | 0 | 1 | 0 | 1 |
| Adverse event related to malaria | 8 | 2 | 1 | 12 | 15 | 3 | 41 |

<sup>a</sup> No adverse events met the criteria for a serious adverse event, resulted in study discontinuation, or were considered related to tafenoquine. <sup>b</sup> The severity of adverse events was assessed in accordance with the Common Terminology Criteria for Adverse Events (26) (mild=grade 1; moderate=grade 2; severe=grade 3; life-threatening consequences=grade 4; death related to AE=grade 5). No grade 4 or 5 adverse events were recorded in this study.

## DISCUSSION

The objective of this study was to characterize the transmission blocking activity of a low dose of tafenoquine in healthy volunteers using the IBSM-transmission model. At the time of study design, the transmission blocking activity of tafenoquine in humans had not previously been assessed. However, concurrently with our trial, another group has assessed the transmission blocking activity of low dose tafenoquine in a field trial in Mali, Africa (27). This presents a unique opportunity to compare the results obtained from a volunteer infection study enrolling a small number of healthy volunteers (*n*=6), with a larger field trial enrolling patients with asymptomatic *P. falciparum* (*n*=80 across 4 tafenoquine dose groups).

The performance of the *P. falciparum* IBSM-transmission model in enabling a robust assessment of a candidate transmission blocking intervention was confirmed in the current study. Asexual parasitemia was successfully cleared in all participants using a piperaquine monotherapy dosing strategy, which allowed for subsequent development of mature transmissible gametocytes. As expected, gametocyte densities (median 234 gametocytes/mL on day 25) were considerably lower than are typically observed in the field (approximately 40,000 gametocytes/mL at baseline in the Mali trial (27)). Nevertheless, all participants were infective to mosquitoes prior to administration of tafenoquine on day 25. Further, a high intensity of mosquito infectivity was observed at this time point, with infectivity rates ≥74% (oocysts) or ≥40% (sporozoites) in 5/6 participants. One participant exhibited a somewhat lower intensity of mosquito infectivity at baseline (22% for oocysts and 4% for sporozoites) compared with the other participants, despite having similar gametocyte levels. Interestingly, this participant had an elevated CRP (46 mg/L at day 25) compared to the other participants (all ≤16 mg/L). An inhibitory effect of inflammatory cytokines on gametocyte transmissibility has been demonstrated in vitro and in an animal model (28, 29), and it is therefore possible that this may have contributed to lower transmission in this participant.The generally high transmission rates achieved at baseline (with relatively low gametocyte densities) were likely facilitated by the Percoll enrichment step prior to mosquito feeding, as well as optimization of the timing of feeding assays relative to the entry of mature gametocytes into the circulation.

The dose of tafenoquine assessed for transmission blocking activity in this study (50 mg single dose) is considerably lower than is used for currently approved indications in individuals with sufficient G6PD activity. Recommended dosing for malaria prophylaxis is 200 mg daily for 3 days (loading) followed by 200 mg weekly (maintenance), while a single 300 mg dose is recommended for radical cure of *P. vivax*. We assessed a dose of 50 mg hypothesizing that such a dose may be safe to administer to individuals of unknown G6PD status, in a similar manner to low dose primaquine. A dose of 100 mg tafenoquine was found to be safe in a small group of G6PD heterozygous females with enzyme activity 40-60% of normal (18). Nevertheless, further work is required to determine the appropriate dose of tafenoquine that could safely be used in the absence of G6PD screening. As expected, no safety signals were associated with tafenoquine dosing of volunteers with G6PD levels in the normal range in the current study. A mild decrease in hemoglobin was observed in all participants over the course of the study relative to malaria infection, consistent with what has been observed in previous IBSM studies (30). Other adverse events recorded during the study were commensurate with blood stage challenge of healthy malaria naïve volunteers. Overall, no significant safety concerns were identified in this study.

A single dose of 50 mg tafenoquine was found to significantly reduce but not abrogate transmission of parasites to *Anopheles* mosquitoes, although a delay in activity was evident. No attenuation of transmission was detected one day after dosing, but a significant reduction in the intensity of mosquito infection was evident 4 days after dosing, with a further reduction observed at 7 days after dosing (median 81% reduction for oocysts and 77% reduction for sporozoites at 7 days post-dose compared to baseline). There was no significant decrease in gametocyte density detected over the course of 7 days post-dose, likely indicating that sterilization of the gametocytes preceded their clearance. A similar delay in tafenoquine transmission blocking activity was observed in the field (27). In the Mali trial, participants received standard treatment with dihydoartemisinin-piperaquine (DP) over 3 days, or DP plus a single dose of 0.42 mg/kg, 0.83 mg/kg, or 1.66 mg/kg tafenoquine. For comparison in our study where participants were not dosed by body weight, the 50 mg tafenoquine dose equates to a body weight adjusted dose range of 0.48-0.88 mg/kg. Two days after the commencement of treatment in the Mali trial, no difference in the reduction in mosquito infection rate was detected between the groups receiving tafenoquine compared to the DP control group. However, a significant difference was observed at 7 days after treatment initiation, with a median 100% reduction (at the oocyst level) in all three tafenoquine dose groups. This reduction is higher than we observed in our study at 7 days post tafenoquine dose (81% at the oocyst level). We hypothesize that this difference may be due to a contribution from the combined DP treatment in the Mali study given that a significant reduction in the mosquito infection rate occurred in the DP control group.

Our study had several limitations. First, due to budget constraints our sample size was small, limiting the accuracy of estimation of the transmission blocking effect. Second, we did not include control participants not receiving tafenoquine, and it is possible that a decline in transmission may have occurred even in the absence of tafenoquine. Finally, we were only able to test a single dose of tafenoquine (50 mg). Testing the effect of other doses would have provided valuable data for identifying a dose-response relationship in transmission blocking activity.

In conclusion, this study has demonstrated the utility of single low dose tafenoquine in reducing transmission of *P. falciparum* from humans to mosquitoes. The results obtained in this volunteer infection study compare favorably with those of a recent field study assessing the transmission blocking activity of low dose tafenoquine in African patients with asymptomatic *P. falciparum* infection. The reason for the delay in the activity of tafenoquine identified in both studies is unclear. The mode of action of the drug and/or a requirement for metabolism-dependent tafenoquine activation may be important factors. Although the delay in the activity of tafenoquine is a drawback compared to its fast acting analogue primaquine, the considerably longer half-life of tafenoquine suggests that it may offer a much longer duration of transmission blocking activity. Such a property would be highly advantageous in preventing transmission of gametocytes that appear in circulation after dosing.

## METHODS

### Study design and participants

This phase 1b study was planned to be composed of three parts. Part 1, performed as planned, was designed to evaluate the potential of single oral doses of tafenoquine to clear asexual blood stage *P. falciparum*. The design and results of this part will be presented elsewhere. Part 2 was designed to evaluate the chemoprophylaxis potential of a single oral dose of tafenoquine. This part was optional and was dependent on the results obtained in Part 1. Following review of the results obtained in part 1 it was decided not to conduct part 2; the study design and methodology of part 2 is not described here. Part 3, performed as planned, was designed to determine if a single oral dose of tafenoquine is active against mature gametocytes and is able to block transmission to mosquitoes. The study design and methodology of part 3 are described below.

This study was an open label, non-randomized, clinical trial using the IBSM model. Healthy malaria naïve males and females (non-pregnant, non-lactating) aged 18-55 years were eligible for inclusion in the study (see Text S1, supplementary file page 5 for full eligibility criteria). The study was conducted at the University of the Sunshine Coast Clinical Trials Unit (Morayfield, Australia) and was registered on the Australian and New Zealand Clinical Trials Registry (Trial ID: ACTRN12620000995976).

### Procedures

The study was conducted in a single cohort of 6 participants. Participants were inoculated intravenously with approximately 2800 viable *P. falciparum* 3D7 infected erythrocytes on day 0. Parasitemia was monitored throughout the study by quantitative PCR (qPCR) targeting the gene encoding *P. falciparum* 18S rRNA (31). Additional blood samples were collected at select time points for quantitative reverse transcriptase PCR (qRT-PCR) targeting male and female gametocyte-specific mRNA transcripts, *pfs25* and *pfMGET* respectively (32), as well as the ring-stage trophozoite-specific transcript *SBP1* (33). Participants were administered piperaquine phosphate tablets (PCI Pharma Services) on day 9 (single 480 mg dose) and day 11 (single 960 mg dose) to clear asexual parasites while allowing development of transmissible gametocytes (24, 25). Transmission of gametocytes to mosquitoes was assessed on day 25 immediately prior to administration of a single oral dose of 50 mg tafenoquine succinate in tablet form (Kodatef^®^, Biocelect). A validated pharmacy method for extemporaneous preparation was used to prepare the dose from the 100 mg tablets supplied by the manufacturer. Additional transmission assessments were performed on days 26, 29 and 32. Participants received a standard curative course of artemether-lumefantrine (Riamet^®^, Novartis Pharmaceuticals) following the final transmission assays on day 32. The end of study occurred on day 35.

Transmission of gametocytes to *Anopheles* mosquitoes was determined by collecting blood samples (up to 66 mL) from participants and conducting enriched membrane feeding assays (eMFAs) as previously described (24). Briefly, blood samples collected at the clinic were transported to QIMR Berghofer Medical Research Institute (under controlled temperature) where eMFAs were conducted. Blood was leukodepleted and uninfected erythrocytes separated from gametocytes using a concentration method (65% Percoll density gradient). The resulting gametocyte rich band was added to malaria naïve donor serum and malaria naïve donor blood to form the mosquito feed mix. Mosquitoes that had been reared in a controlled environment in a PC3 insectary (BIC3) were distributed into containers with gauze lids and starved prior to feeding. Two separate pots containing approximately 75 mosquitoes each were allowed to feed for approximately 30 minutes through parafilm membranes on water jacketed glass feeders attached to a pre-warmed water bath (39-40°C). After feeding, the number of engorged mosquitoes was recorded. The blood-fed mosquitoes were maintained on glucose in a controlled environment (27°C, 70-80% relative humidity). The mosquitoes from one pot were dissected after 8 days, with approximately 50 midgut dissections analyzed for oocysts by 18S qPCR (34) and the remaining midguts examined by microscopy for visual confirmation of oocysts. The mosquitoes from the other pot were dissected after 17 days, with approximately 50 head/thorax dissections analyzed for sporozoites by 18S qPCR (see Text S2, supplementary file page 9).

Venous blood samples (2 mL) were collected at days 25, 26, 27, 29 and 32 for the measurement of plasma tafenoquine concentrations by LC-MS/MS. Ethylenediaminetetraacetic acid was used as the anticoagulant. All blood samples were centrifuged at 1600*g* for 15 minutes at 6±1°C, and the separated plasma samples were stored at -80° ± 5°C until analysis. Plasma concentrations of tafenoquine were measured using a validated LC-MS/MS method. Briefly, plasma samples (50 µL) were precipitated by the addition of 200 μL acetonitrile containing a stable-isotope label tafenoquine as internal standard. Extracts were separated on a Waters Atlantis T3 column. (3 μm) with a linear 0.1% formic acid, acetonitrile gradient and injected into a Sciex Qtrap 4000 mass spectrometer in the positive MRM mode. Unknown sample MSMS peak areas were integrated and read off a 6-point calibration curve (range 1 to 1200 ng/mL) with a quadratic regression analysis and weighting of 1/x^2^. A correlation coefficient greater than or equal to 0.9970 was obtained for all calibration curves. The intra- and inter-assay precisions (coefficient of variation %) were 0.5% (*n*=6) and 4.4% (*n*=8), respectively, for tafenoquine at a limit of quantification of 1 ng/mL. Corresponding values were 1.6% and 3.2% at 100 ng/mL. The mean ± SD inter-assay accuracy at 1 and 100 ng/mL were 102.0 ± 4.5% and 100.9 ± 3.3%, respectively.

Safety assessments included adverse event (AE) recording, clinical laboratory parameters (hematology, biochemistry and urinalysis), vital signs (blood pressure, heart rate, respiratory rate, and body temperature), 12-lead electrocardiography, and physical examination. AEs were recorded from the time of inoculation with the malaria challenge agent up to the end of the study. AE severity was recorded in accordance with the Common Terminology Criteria for Adverse Events (26) (mild=grade 1; moderate=grade 2; severe=grade 3; life-threatening consequences=grade 4; death related to AE=grade 5). In addition, an AE was classified as a serious adverse event (SAE) if it met one of the following criteria: resulted in death, was life-threatening, required inpatient hospitalization, resulted in persistent or significant disability, was a congenital anomaly, was considered medically important, or constituted a possible Hy’s Law case. The investigator assessed whether AEs were related to tafenoquine, piperaquine, and/or to the malaria challenge agent.

## Outcomes

The primary outcome of this clinical trial was associated with part 1 and will be presented elsewhere. Secondary outcomes associated with part 3 of the trial were the reduction in gametocytemia and mosquito infection on days 26, 29 and 32 compared to day 25; and the incidence, severity, and relationship to tafenoquine of AEs, and change from baseline in clinical laboratory parameters (hematology, biochemistry, urinalysis).

### Statistical analysis

We had initially intended to enroll a total of 14 participants, based on the assumption that 75% of participants would be infective at baseline. With 14 participants, there would be 10 participants infective at day 25 (lower bound of 95% CI of 41.9%). Of those 10 participants infective at day 25, assuming a 95% transmission blocking efficacy for tafenoquine treatment, then the lower bound of the 95% CI for the reduction in mosquito infectivity would be 69.2%. Due to budget constraints implemented part-way through the study, only 6 participants could be enrolled.

The change in transmission to mosquitoes on days 26, 29 and 32 compared to day 25 (pre-tafenoquine dosing) was reported as the proportion of participants infective to mosquitoes, and the intensity of mosquito infection (proportion of infected mosquitoes), at pre- and post-tafenoquine treatment time points. The median within participant percentage reduction in the proportion of infected mosquitoes relative to the pre-tafenoquine time point (baseline) was calculated. Separate analyses were performed at the oocyst and sporozoite levels. A participant was defined as infective to mosquitoes if at least one mosquito was positive for the presence of oocysts or sporozoites. A mosquito was defined as infected if the presence of oocysts or sporozoites were detected in midgut or head/thorax dissections respectively by 18S qPCR. Changes in the proportion of participants infective to mosquitoes between day 25 and each of the post-tafenoquine treatment time points were assessed using McNemar’s test. A Wilcoxon signed-rank test was used to assess whether there was a nonzero percentage reduction in infected mosquitos between baseline (pre-tafenoquine) and post-treatment time points.

Spearman’s rank order correlation was used to assess factors associated with transmission at day 25 (pre-tafenoquine baseline). Repeated measures correlation was run for assessment of oocyst vs sporozoite infectivity, and for qPCR vs microscopy oocyst infectivity, following logit(x+0.01) transformations, to account for multiple measures per participant across all transmission days (35, 36). The area under the parasitemia-time curve prior to piperaquine dosing (day 4 to day 9) was calculated on the parasites/mL scale for each participant using the trapezoidal rule method.

Gametocyte densities were quantified using 18S qPCR, as it was confirmed during the study (by qRT-PCR targeting *SBP1* mRNA) that asexual parasites were successfully cleared following piperaquine dosing and thus the 18S signal detected during the transmission phase of the study represented gametocytes only. A Wilcoxon signed-rank test was used to assess whether there was a nonzero percentage reduction in gametocyte density between baseline (pre-tafenoquine) and post-treatment time points. The change in the ratio of *pfMGET* transcript levels to *pfs25* transcript levels on days 26, 29 and 32 compared with day 25 (pre-tafenoquine baseline) was assessed using a Wilcoxon rank sum test.

Statistical significance was set at a *P*-value < 0.05 (two-sided). Statistical analysis were performed in Stata version 15 (StataCorp, College Station, TX, USA) and in R statistical package version 4.1.0 (R Core Team, 2020).

### Study approval

This study was approved by the QIMR Berghofer Medical Research Institute Human Research Ethics Committee (reference number P3646) with mutual recognition of the ethical review of the QIMR Berghofer HREC P3646 by the Departments of Defence and Veterans’ Affairs Human Research Ethics Committee (DDVA HREC 194-19). All participants gave written informed consent before enrollment.

## Data Availability

All data produced in the present study are available upon reasonable request to the authors.

## AUTHOR CONTRIBUTIONS

Study conceptualization:BEB, JSM, GDS.

Study design: BEB, JSM, GDS, LM, SLL, RW, AJP, HJ.

Study management: RW, BEB.

Data acquisition: HM, JP, JH, BON, JG, SL, FA, ME, GB, BEB.

Data analysis: SLL, LM.

Manuscript writing: AJP, BEB. Manuscript review: All authors.

## ACKNOWLEWDGEMENTS

We thank all the volunteers who participated in the study; staff at the University of the Sunshine Coast Clinical Trials Unit who conducted the trial; staff at the Queensland Paediatric Infectious Diseases laboratory for qPCR analysis; Dr David Wesche (Certara), Dr Jörg Möhrle (Medicine for Malaria Venture), and Dr Scott Miller (Bill and Melinda Gates Foundation) for their participation in the Safety Data Review Team; Karin Van Breda for the measurement of plasma tafenoquine concentrations using LC-MS/MS; and Medicines for Malaria Venture (MMV) for providing financial support to the Clinical Malaria Group within QIMR Berghofer Medical Research Institute.

## DISCLAIMER

The views expressed in this article are those of the authors and do not necessarily reflect the official policy or position of the Australian Defence Force, Joint Health Command or any extant Australian Defence Force policy.

## SUPPLEMENTARY MATERIAL

- **Figure S1**. Individual participant (A) and male (B) gametocytemia-time profiles.
- **Figure S2**. Individual participant hemoglobin-time profiles.
- **Table S1**. Incidence of adverse events by participant.
- **Text S1**. Participant eligibility criteria.
- **Text S2**. qPCR analysis of mosquito head/thorax dissections.

