## Supplementary material for "Transmission blocking activity of low dose tafenoquine in healthy volunteers experimentally infected with *Plasmodium falciparum*"

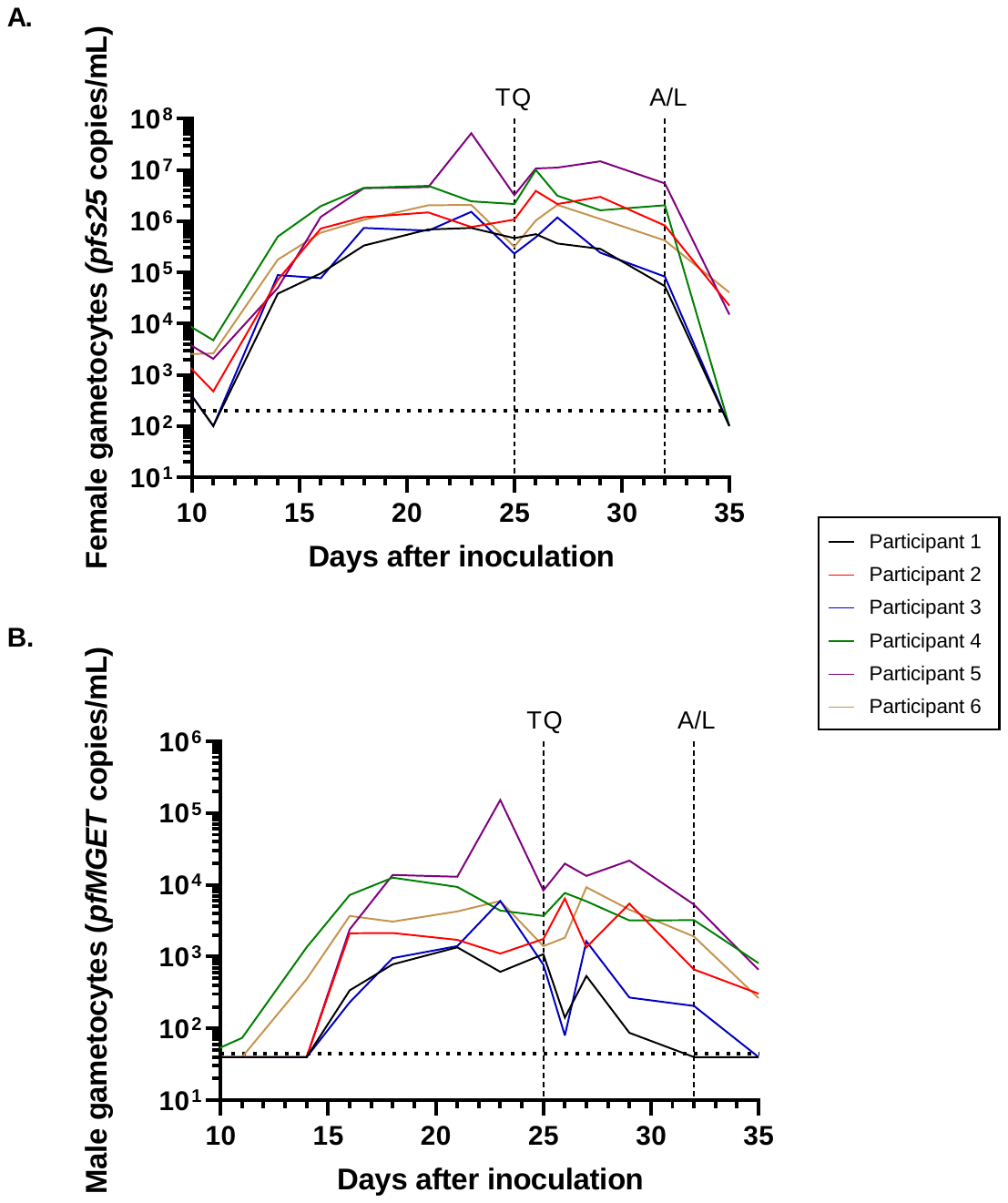

**Figure S1.** Individual participant female (A) and male (B) gametocytemia-time profiles. Participants were inoculated intravenously with *P. falciparum*-infected erythrocytes on day 0 and were administered an oral dose of piperaquine (PPQ) on day 9 (480 mg) and day 11 (960 mg) to clear asexual parasitaemia while allowing transmissible gametocytes to mature. A single oral dose of 50 mg tafenoquine (TQ) was administered on day 25. Definitive antimalarial treatment with a standard 3-day course of artemether-lumefantrine (A/L) was initiated on day 32. Female gametocytemia was detected using qRT-PCR targeting the *pfs25* transcript while male gametocytemia was detected using qRT-PCR targeting the *pfMGET* transcript. The horizontal dotted lines represent the limit of detection of the qRT-PCR assays.

**Table S1.** Change in the ratio of pfMGET to pfs25 transcripts from pre-tafenoquine dosing to post-treatment time-points.

| **Time-point (days relative to tafenoquine dosing)** | **N** | **Median transcript ratio (range)** | **Median change in transcript ratio (range)** | **z** | ***P*-value*** |
| --- | --- | --- | --- | --- | --- |
| 0 (pre-tafenoquine) | 6 | 0.0022 (0.0017 – 0.0044) | **-** | **-** | **-** |
| 1 | 6 | 0.0012 (0.0001 – 0.0019) | -0.0013 (-0.0034 – 0.0) | 1.99 | 0.046 |
| 4 | 6 | 0.0017 (0.0003 – 0.0040) | -0.0007 (-0.0023 – 0.0003) | 1.57 | 0.12 |
| 7 | 6^#^ | 0.0018 (0.0008 – 0.0045) | -0.0005 (-0.0016 – 0.0003) | 1.15 | 0.25 |

^#^ *pfMGET* transcripts could not be detected in one participant at the 7 day post-doing time point. For the purpose of analysis, a value corresponding with half the limit of detection of the assay was assigned. *Change in transcript ratio was assessed using a Wilcoxon rank sum test.

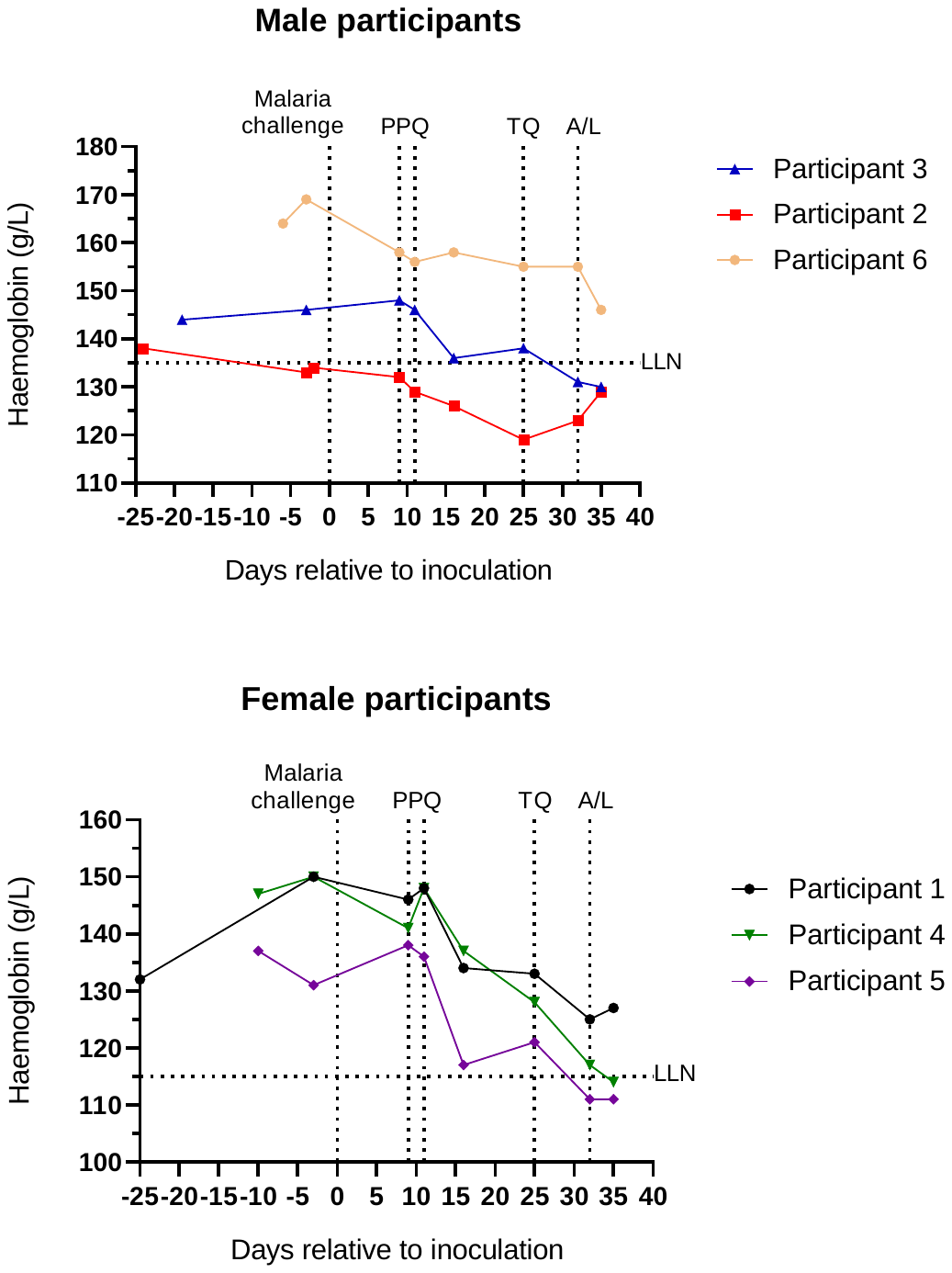

### **Figure S2.** Individual participant hemoglobin-time profiles.

LLN= lower limit of normal.

### **Table S1.** Incidence of adverse events by participant.

| **Adverse Event Preferred Term** | **Number of events** | | | | | | |
| --- | --- | --- | --- | --- | --- | --- | --- |
|  | **Participant 1** | **Participant 2** | **Participant 3** | **Participant 4** | **Participant 5** | **Participant 6** | **Total** |
| Arthralgia | 1 |  |  |  |  |  | 1 |
| Back pain | 1 |  |  |  | 2 |  | 3 |
| Chills |  |  |  | 2 | 1 |  | 3 |
| Decreased appetite |  |  |  |  | 2^b^ |  | 2 |
| Eye pain | 1 |  |  |  |  |  | 1 |
| Fatigue |  |  |  |  | 1^a^ |  | 1 |
| Groin pain |  | 1^a^ |  |  |  |  | 1 |
| Hemoglobin decreased | 1 |  |  | 1 |  | 1 | 3 |
| Headache | 1 | 1 | 2 | 4 | 5^c^ | 2 | 15 |
| Hot flush |  |  |  |  | 1 |  | 1 |
| Hyperhidrosis |  | 2 |  |  |  |  | 2 |
| Influenza like illness |  |  |  | 1 |  |  | 1 |
| Lethargy |  |  | 1 |  |  |  | 1 |
| Lymphocyte count decreased |  |  | 1^a^ | 1^a^ | 1^d^ |  | 3 |
| Musculoskeletal pain |  | 1 |  |  |  |  | 1 |
| Myalgia |  |  |  |  | 1 |  | 1 |
| Nausea | 1 |  | 1 |  | 3^a^ |  | 5 |
| Neck pain | 1 |  |  |  |  |  | 1 |
| Neutrophil count decreased | 1^a^ |  |  |  |  |  | 1 |
| Oropharyngeal pain |  |  |  |  | 1^a^ |  | 1 |
| Pyrexia |  |  |  | 2 | 1 |  | 3 |
| Rash |  |  |  | 1 |  |  | 1 |
| Sinus tachycardia |  |  |  | 4 |  |  | 4 |
| Stress |  |  | 1 |  |  |  | 1 |
| Upper respiratory tract infection |  | 1 |  |  |  |  | 1 |
| Vessel puncture site hematoma |  |  | 1 |  |  |  | 1 |
| Vomiting |  |  |  |  | 1 |  | 1 |
| Vulvovaginal pruritus |  |  |  |  | 1 |  | 1 |
| White blood cell count decreased | 1^a^ |  |  | 1^a^ |  |  | 2 |

^a^ Event/s of moderate (grade 2) intensity. ^b^ One of the events moderate (grade 2) in intensity. ^c^ Two of the events moderate (grade 2) in intensity. ^d^ Event of severe intensity.

### **Text S1.** Participant eligibility selection criteria

**Inclusion Criteria**

Volunteers who met all of the following criteria were eligible for inclusion in the study:

**Demography:**

1. Male or female (non-pregnant, non-lactating) aged 18 to 55 years inclusive who was contactable and available for the duration of the study and up to two weeks following the end of study visit.
2. Total body weight greater than or equal to 50 kg, and a body mass index (BMI) within the range of 18 to 32 kg/m^2^ (inclusive). BMI is an estimate of body weight adjusted for height. It is calculated by dividing the weight in kilograms by the square of the height in metres.

**Health status:**

3. Certified as healthy by a comprehensive clinical assessment (detailed medical history, complete physical examination and special investigations).
4. At least normal G6PD enzyme activity levels as defined by the parameters of the specific quantitative G6PD test employed at Screening.
5. Vital signs at screening and pre-inoculation (measured after five minutes in the supine position):
   - Systolic blood pressure (SBP) 90-140 mmHg,
   - Diastolic blood pressure (DBP) 40-90 mmHg,
   - Heart rate (HR) 40-100 bpm.
6. Electrocardiograph (ECG) ranges at screening and pre-inoculation: QTcF ≤450 ms for males, QTcF ≤470 ms for females, and PR interval ≤210 ms.
7. Female volunteers of childbearing potential who had, or may have had, male sexual partner(s) during the course of the study were to use an insertable (implant and IUD, injectable, transdermal or combination oral contraceptive approved by the Australian Therapeutic Goods Administration (TGA) combined with a barrier contraceptive from the day of informed consent through to 90 days after the last dose of tafenoquine. Abstinent females were to agree to start a double method if they started a sexual relationship with a male during the study. Adequate contraception did not apply to volunteers of childbearing potential with same sex partners (abstinence from penile-vaginal intercourse), when this was their preferred and usual lifestyle. Females must not have been planning *in vitro* fertilisation within the required contraception period.

Women of non-childbearing potential who did not require contraception during the study were defined as: surgically sterile (tubal ligation was not considered surgically sterile), post-menopausal (spontaneous amenorrhoea for ≥12 months, or spontaneous amenorrhoea for 6-12 months and follicle-stimulating hormone (FSH) ≥40 IU/mL; either was to be together with the absence of oral contraceptive use for >12 months).

Male volunteers who had, or may have had, female sexual partner(s) during the course of the study were to agree to use a double method of contraception including condom plus diaphragm, or condom plus insertable device, or condom plus stable oral/transdermal/injectable hormonal contraceptive by the female partner, from the time of informed consent through to 90 days after the last dose of tafenoquine. Abstinent males were to start a double method if they started a sexual relationship with a female during the study, and through to 90 days after the last dose of tafenoquine. Male volunteers with female partners that were surgically sterile, or males who had undergone sterilisation and had testing to confirm the success of the sterilisation may have also been included.

**Regulations:**

8. Completion of the written informed consent process prior to undertaking any study-related procedure.
9. Must have been willing and able to communicate and participate in the whole study.
10. Agreement to adhere to the lifestyle considerations specified in the study protocol throughout the study duration.

**Exclusion Criteria**

Volunteers who met any of the following criteria were not eligible for inclusion in the study:

**Medical history and clinical status:**

1. Volunteer who lives alone (at any stage from inoculation day until the end of the study). Volunteers who lived alone may have been included on a case-by-case basis, following discussion with the Principal Investigator and with approval of the Medical Monitor. Volunteers who lived alone must have identified and provided contact details of a support person who was aware of the volunteer’s participation in the study and was available to provide assistance if required (for example with contacting the volunteer in the event that study staff were unable to, or with transporting the volunteer to and from the study site if required).
2. Any history of malaria or participation in a previous malaria challenge study or malaria vaccine study.
3. Must not have travelled to or lived (>2 weeks) in a malaria-endemic region during the past 12 months or planned travel to a malaria-endemic region during the course of the study. Must not have lived for >1 year in a malaria-endemic region in the past 10 years. Must not have ever lived in a malaria-endemic region for more than 10 years inclusive.
4. Had evidence of increased cardiovascular disease risk (defined as >10%, 5-year risk for those greater than 35 years of age, as determined by the Australian Absolute Cardiovascular Disease Risk Calculator (http://www.cvdcheck.org.au/)). Risk factors include sex, age, systolic blood pressure (mm/Hg), smoking status, total and HDL cholesterol (mmol/L), and reported diabetes status.
5. History of splenectomy.
6. Volunteer was unwilling to defer blood donations to the Blood Service for at least six months after the end of study visit.
7. Volunteer who had ever received a blood transfusion.
8. Any recent (<6 weeks) or current systemic therapy with an antibiotic or drug with potential antimalarial activity (e.g. chloroquine, piperaquine phosphate, benzodiazepine, flunarizine, fluoxetine, tetracycline, azithromycin, clindamycin, doxycycline etc.).
9. Known hypersensitivity to artesunate or any of its excipients, artemether or other artemisinin derivatives, proguanil/atovaquone, primaquine, or 4-aminoquinolines.
10. Hematology, clinical chemistry or urinalysis results at screening or at the day -1 to day -3 eligibility visit that were outside of Sponsor-approved clinically acceptable laboratory ranges, and were considered clinically significant by the Investigator.
11. Participation in any investigational product study within the 12 weeks preceding inoculation.
12. Symptomatic postural hypotension at screening (confirmed on two consecutive readings), irrespective of the decrease in blood pressure, or asymptomatic postural hypotension defined as a decrease in systolic blood pressure ≥20 mmHg within 2-3 minutes when changing from supine to standing position.
13. History or presence of diagnosed (by an allergist/immunologist) or treated (by a physician) food or known drug allergies (including but not limited to allergy to any of the antimalarial rescue treatments), or any history of anaphylaxis or other severe allergic reactions including face, mouth, or throat swelling or any difficulty breathing. Volunteers with seasonal allergies/hay fever or allergy to animals or house dust mite that were untreated and asymptomatic at the time of dosing could be enrolled in the study.
14. History of convulsion (including intravenous drug or vaccine-induced episodes). A medical history of a single febrile convulsion during childhood was not an exclusion criterion.
15. Presence of current or suspected serious chronic diseases such as cardiac or autoimmune disease (HIV or other immuno-deficiencies), insulin-dependent and non-insulin dependent diabetes, progressive neurological disease, severe malnutrition, acute or progressive hepatic disease, acute or progressive renal disease, porphyria, psoriasis, rheumatoid arthritis, asthma (excluding childhood asthma, or mild asthma with preventative asthma medication required less than monthly), epilepsy, or obsessive-compulsive disorder.
16. History of malignancy of any organ system (other than localised basal cell carcinoma of the skin or *in situ* cervical cancer), treated or untreated, within five years of screening, regardless of whether there was evidence of local recurrence or metastases.
17. History of schizophrenia, bi-polar disease, psychoses, disorders requiring lithium, attempted or planned suicide, or any other severe (disabling) chronic psychiatric diagnosis.
18. Volunteers who had received psychiatric medications within one year prior to enrolment for a psychiatric condition, or who had been hospitalised within five years prior to enrolment for either a psychiatric illness or due to danger to self or others.
19. History of an episode of minor depression that required at least six months of pharmacological therapy and/or psychotherapy within the last five years; or any episode of major depression.

The Beck Depression Inventory was used as an objective tool for the assessment of depression at screening. In addition to the conditions listed above, volunteers with a score of 20 or more on the Beck Depression Inventory and/or a response of 1, 2 or 3 for item 9 of this inventory (related to suicidal ideation) were not eligible for participation. These volunteers were to be referred to a general practitioner or medical specialist as appropriate. Volunteers with a Beck score of 17 to 19 may have been enrolled at the discretion of the Investigator if they did not have a history of the psychiatric conditions mentioned in this criterion and their mental state was not considered to pose additional risk to the health of the volunteer or to the execution of the study and interpretation of the data gathered.

20. History of recurrent headache (e.g. tension-type, cluster or migraine) with a frequency of ≥2 episodes per month on average and severe enough to require medical therapy, during the 5 years preceding screening.
21. Presence of clinically significant infectious disease or fever (e.g. sublingual temperature ≥38.5°C) within the five days prior to inoculation.
22. Evidence of acute illness within the four weeks prior to screening that the investigator deemed may have compromised volunteer safety.
23. Significant inter-current disease of any type, in particular liver, renal, cardiac, pulmonary, neurologic, rheumatologic, or autoimmune disease by history, physical examination, and/or laboratory studies including urinalysis.
24. Volunteer had a clinically significant disease or any condition or disease that might affect drug absorption, distribution or excretion (e.g. gastrectomy, diarrhoea).
25. Blood donation of any volume or participation in any research study involving blood sampling (more than 450 mL/unit of blood) within one month before tafenoquine administration, or blood donation to Australian Red Cross Blood Service (Blood Service) or other blood bank during the eight weeks prior to inoculation.
26. Medical requirement for intravenous immunoglobulin or blood transfusions.
27. History or presence of alcohol abuse (alcohol consumption more than 40 g/4 units/4 standard drinks per day), or drug habituation, or any prior intravenous usage of an illicit substance.
28. Female volunteer who was breastfeeding.
29. Any vaccination within the last 28 days.
30. Any corticosteroids, anti-inflammatory drugs (excluding commonly used over-the-counter anti-inflammatory drugs such as ibuprofen, acetylsalicylic acid, diclofenac), immunomodulators or anticoagulants within the past three months. Any volunteer currently receiving or having previously received immunosuppressive therapy (including systemic steroids, adrenocorticotrophic hormone or inhaled steroids) at a dose or duration potentially associated with hypothalamic-pituitary-adrenal axis suppression within the past year.
31. Use of prescription drugs (excluding contraceptives) or non-prescription drugs or herbal supplements (such as St John’s Wort), within 14 days or five half-lives (whichever was longer) prior to inoculation. Limited use of other non-prescription medications or dietary supplements, not believed to affect volunteer safety or the overall results of the study, may have been permitted on a case-by-case basis following approval by the sponsor in consultation with the investigator. Volunteers were requested to refrain from taking non-approved concomitant medications from recruitment until the conclusion of the study.
32. Cardiac/QT risk:

- Family history of sudden death or of congenital prolongation of the QTc interval or known congenital prolongation of the QTc interval or any clinical condition known to prolong the QTc interval.
- History of symptomatic cardiac arrhythmias or with clinically relevant bradycardia.
- Electrolyte disturbances, particularly hypokalaemia, hypocalcaemia, or hypomagnesaemia.
- ECG abnormalities in the standard 12-lead ECG (at screening and prior to inoculation) which in the opinion of the Investigator was clinically relevant or would interfere with the ECG analyses.

33. History of retinal abnormalities, diseases of the retina or macula of the eye, visual field defects, and hearing disorders like reduced hearing and tinnitus.

**General conditions:**

34. Any volunteer who, in the judgment of the investigator, was likely to be non-compliant during the study, or was unable to cooperate because of a language problem or poor mental development.
35. Any volunteer in the exclusion period of a previous study according to applicable regulations.
36. Any volunteer who was the investigator, research assistant, pharmacist, study coordinator, or other staff thereof, directly involved in conducting the study.
37. Any volunteer without good peripheral venous access.

**Biological status:**

38. Positive result on any of the following tests: hepatitis B surface antigen (HBs Ag), anti-hepatitis B core antibodies (anti-HBc Ab), anti-hepatitis C virus (anti-HCV) antibodies, anti-human immunodeficiency virus 1 and 2 antibodies (anti-HIV1 and anti-HIV2 Ab).
39. Positive urine drug test. Any drug in the urine drug screen unless there was an explanation acceptable to the investigator (e.g., the volunteer had stated in advance that they had consumed a prescription or over-the-counter product which contained the detected drug) and/or the volunteer had a negative urine drug screen on retest by the pathology laboratory. Any volunteer testing positive for acetaminophen (paracetamol) at screening and/or inoculation day may still have been eligible for study participation, at the investigator’s discretion.
40. Positive alcohol breath test.

### **Text S2.** qPCR analysis of mosquito head/thorax dissections

DNA extraction

Mosquito head/thorax dissections were collected in 180 μl ATL buffer (Qiagen) and 5 μl DNA extraction control (DEC) was added prior to processing. Bead beating was performed using a Mini-Beadbeater (Sigma-Aldrich) for 2 minutes with 2.3 mm Zirconia/Silica beads. 40 μl Proteinase K (Qiagen) was added and the samples were incubated for a minimum of 3 hours. DNA extraction was performed using a DNeasy Blood and Tissue Kit (Qiagen) in accordance with the manufacturer’s instructions. DNA was eluted in 50 μl AE buffer (Qiagen).

qPCR assays

qPCR assays were performed using a QuantiNova Probe PCR Kit (Qiagen) and Bio-Rad CFX384 Real-Time PCR Detection System in accordance with the manufacturers’ instructions. Details of primers and probes targeting the gene encoding *Plasmodium falciparum* 18S rRNA (SAF18S assay [1]), the gene encoding *Anopheles* ribosomal protein S7 (RPS7 assay; mosquito tissue control), and the DEC assay are as follows:

| **Assay** | **Primer Sequences** | **Probe Sequence** |
| --- | --- | --- |
| **SAF18S** | Forward  5’-AGGAAGTTTAAGGCAACAACAGGT-3’ | 5’- /5Cy5/TGTCCTTAGATGAACTA  GGCTGCACGCG/3IAbRQSp/ -3’ |
|  | Reverse  5’-GCAATAATCTATCCCCATCACGA-3’ |  |
| **RPS7** | Forward  5’-TGGAAATGAACTCGGATCTGAAG-3’ | 5’- /5HEX/CAGCTGCGT/ZEN/GATCTG  TACATCACCCGCGC/3IABkFQ/ -3’ |
|  | Reverse  5’-CCTTCTTGTTGTTGAACTCGACCT-3’ |  |
| **DEC** | Manufacturer Supplied | |

Standard curve and control preparation

An 18S rRNA qPCR standard curve was generated using serial dilutions of a synthetic plasmid DNA standard. The DNA standard was prepared by PCR amplification of *P. falciparum* genomic DNA using SAF18S primers (generating a 199 bp fragment), cloning of PCR fragments into the pGEM-T vector (Promega), and subsequent transformation into *E. coli* using standard laboratory procedures. Recombinant clones were confirmed by Sanger sequencing, linearized and quantitated using a Qubit Fluorometer (Thermo Fisher).

Positive mosquito controls were prepared using naïve blood fed mosquitos. Positive parasite controls were prepared using parasite infected red blood cells at 5% parasitemia. Negative controls were prepared using naïve human blood. DNA extraction of controls was performed as described above.

18S rRNA qPCR assay performance specifications

| **Reportable Range:** | 2.0×10^6^ to 2.0×10^1^ copies/µL |
| --- | --- |
| **Analytical Sensitivity:** |  |
| Limit of detection (LoD) | 3.4 copies/µL |
| Lower Limit of quantification (LLoQ)  Upper Limit of quantification (ULoQ) | 3.4 copies/µL  2.0×10^6^ copies/µL |
| **Precision:** |  |
| Intra-assay variability | %CV = 2.24 |
| Inter-assay variability | %CV = 2.21 |
| **Specificity:** |  |
| Analytical specificity | 100% |
| **Accuracy (Trueness):** |  |
| Comparison-of-methods study  Microscopy versus qPCR | % Agreement = 90%  3 samples were qPCR positive and microscopy negative |
| **Reference Interval:** | Not detect |

1. McCarthy et al. 2013. Experimentally Induced Blood-Stage *Plasmodium vivax* Infection in Healthy Volunteers. J Infect Dis. 208: 1688-1694.
